# Minding the Gap in Sentinel Surveillance Networks: An Analysis of Brazilian Indigenous Areas

**DOI:** 10.1101/2025.02.10.25321997

**Authors:** Juliane F. Oliveira, Adriano O. Vasconcelos, Andrêza L. Alencar, Maria Célia L. S. Cunha, Izabel Marcilio, Manoel Barral-Netto, Pablo Ivan P. Ramos

**Affiliations:** Center for Data and Knowledge Integration for Health (CIDACS), Gonçalo Moniz Institute, Oswaldo Cruz Foundation (FIOCRUZ); Salvador, 41745-715, Brazil; Centre of Mathematics of the University of Porto (CMUP), Department of Mathematics, Porto, Portugal; Luiz Coimbra Institute of Graduate and Engineering Research (COPPE), Federal University of Rio de Janeiro (UFRJ); Rio de Janeiro, 21941-859, Brazil; Department of Computer Science, Federal Rural University of Pernambuco; Recife, 52171-900, Brazil; Bahiana School of Medicine and Public Health (EBMSP); Salvador, 40290-000, Brazil; Medicine and Precision Public Health Laboratory (MeSP^2^), Gonçalo Moniz Institute, Oswaldo Cruz Foundation (FIOCRUZ); Salvador, 40296-710, Brazil

**Keywords:** Representative sentinel surveillance, early pathogen detection, Indigenous health, human mobility, surveillance network optimization, infectious disease surveillance, public health strategy, Brazil

## Abstract

**Background:** Optimizing the allocation of sentinel surveillance sites, particularly aiming early pathogen detection, remains a persistent challenge. Equally critical is ensuring the inclusion of vulnerable and often underserved populations as targets when prioritizing surveillance sites.

**Objective:** This study assesses the current coverage of the respiratory pathogen surveillance network in Brazil and proposes an optimized rearrangement of sentinel locations that balance the coverage of Indigenous populations while integrating country-wide human mobility patterns.

**Methods:** We collected the locations of Indigenous populations living in Special Indigenous Sanitary Districts (DSEIs) from the Brazilian Ministry of Health, and derived mobility route estimates using the Ford-Fulkerson algorithm applied to input air, road, and water transportation data for the whole country. To optimize sentinel city selection for sample collection, we applied a linear optimization algorithm designed to maximize two key objectives: 1) representation of Indigenous regions and 2) coverage of human mobility patterns, thereby enhancing early pathogen detection. Validation of the strategy was performed by obtaining the current list of cities in Brazil’s influenza sentinel network combined with a health attraction index from the Brazilian Institute of Geography and Statistics, which enabled us to assess the suitability and potential benefits of our optimized surveillance network.

**Results:** Our optimization model provides actionable recommendations for pathogen sampling locations, enhancing early detection. By selecting 199 cities, we create a more representative sentinel network, covering all DSEIs by rearranging 108 cities (58.3%), addressing gaps in 9 of 34 previously uncovered regions. This improves nationwide mobility coverage by 16.8 percentage points—from 52.4% to 69.2%—compared to Brazil’s current sentinel system. Additionally, all newly selected cities serve as hubs for medium to high-complexity healthcare, with 37.9% of DSEI cities lacking coverage in the existing flu sentinel network.

**Conclusions:** We propose a strategic framework for sentinel site placement that maximizes DSEI coverage and aligns with human mobility patterns across Brazil, improving the overall effectiveness of disease surveillance, particularly in these critical regions and often underserved populations.

## Introduction

Sentinel surveillance, which involves systematic and regular clinical sample collection to monitor the emergence of infectious diseases across a network of sites, is a key component of public health strategy [1]. Ideally, this network should be representative of the general population while also prioritizing high-risk subpopulations, such as Indigenous groups [2, 3, 4].

Despite decreasing genomic sequencing costs and advancements in metagenomics that allow for the monitoring of multiple pathogens and the identification of novel agents, low- and middle-income countries face challenges when scaling up sentinel surveillance due to limited financial resources [5 - 8]. As a result, sentinel sites are often chosen based on convenience, leaving minority populations, such as Indigenous communities, underserved. In this context, the use of alternative data streams to understand pathogen emergence and the likely pathway of disease spread may maximize the available information for improving data-driven allocation of surveillance resources [6 - 8].

The boundaries between wild landscapes and human settlements are increasingly blurred by climate change, agricultural and urban expansion, deforestation, and landscape fragmentation. These compounding factors heighten the vulnerability of Indigenous communities to infectious disease spillover, which can rapidly spread to urban areas [9]. In addition, Brazilian Indigenous populations face significant barriers to healthcare access, limiting their awareness of circulating diseases and disproportionately increasing their disease burden [2 - 4, 10]. During the COVID-19 pandemic, the incidence and mortality from the disease among Indigenous and Traditional People were higher compared to the general population, highlighting the need for tailored public health strategies to address health disparities in these communities [11 - 14].

We recently showed how human mobility patterns can inform the redesign of current sentinel networks to improve early pathogen detection [8]. However, the high risk of pathogen emergence in Indigenous communities may remain undetected if relying solely on mobility data, potentially delaying the identification of health threats in these populations. To address this gap, we built and expanded on our mobility-based model by explicitly incorporating the geographical distribution of Indigenous communities in Brazil. This enabled the construction of an optimized list of cities that could serve as candidates for early pathogen detection in the country, while also adding an equity component into sentinel planning.

## Methods

### Data sources

For our analyses, we collected data on cities within Brazilian Indigenous Health Districts (DSEI, from the Portuguese *Distrito Sanitário Especial Indígena*) obtained from the Brazilian Ministry of Health, along with mobility coverage data derived from the Ford-Fulkerson algorithm [8, 15].

To ensure the practical applicability of our results, we also collected information on the composition of Brazil’s current influenza sentinel surveillance network, obtained through direct communication with the Ministry of Health. We also incorporated a health attraction index, which quantifies the potential of a city to attract individuals seeking healthcare services. This index, estimated by the Brazilian Institute of Geography and Statistics [16], categorizes cities based on their capacity to provide low-, medium-, and high-complexity health services, further refining our sentinel site selection process. The complete dataset is available in our GitHub directory [17].

### Optimization Problem

To identify the most suitable sentinel sites maximizing both human mobility coverage as well as the representation of DSEIs, we applied an optimization approach.

Brazil’s 34 DSEIs are administrative regions designed to manage health activities for Indigenous territories in an ethnically and culturally sensitive manner. DSEIs encompass 1,470 of Brazil’s 5,570 municipalities, with partial overlap in 46 others, as Indigenous territories do not always align with political boundaries. In our work, we considered a DSEI to be covered by a sentinel network if it includes at least one city which is fully within a DSEI.

To define mobility coverage, we constructed an intercity mobility network, represented as a graph G = (V, E), where the set of nodes V corresponds to cities and the set of links E represents movement frequencies between them. Using this network, we applied the Ford-Fulkerson algorithm [8] to model potential disease spread pathways, identifying the most probable routes for pathogen transmission from a source city. This approach allowed us to determine key locations for early detection by analyzing how frequently cities appeared at different stages in these transmission paths. Further methodological details are available in [8]. Based on the output of the Ford-Fulkerson algorithm, we defined the mobility coverage of a city A over a city B as follows: city A covers city B if n% of the most likely transmission paths originating from city B include city A as their first step, where n is a value between 0 and 100. When n = 100, we concluded that city A fully covers city B.

To maximize mobility coverage while ensuring that all DSEI regions are represented, we formulate this as an optimization problem. Mathematically, let *x*_*i*_ be a binary variable, where *x*_*i*_ = 1 if city *i* is selected for the sentinel network and *x*_*i*_ = 0 otherwise. Let ω_*i*_ represent the mobility coverage weight of city *i*. Additionally, let *R*_*j*_ denote the set of cities within DSEI region *j* and let *N* be the total number of cities to be selected for the sentinel network. The optimization function is then defined as:

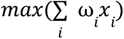

subject to

1. 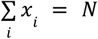, that is, select N cities to compose the sentinel.
2. 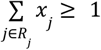 for all *j* ∈ {1, 2, …, 34}, that is, for each DSEI region *j*, at least one city in *R*_*j*_ is selected.

The optimization problem is implemented in Python using PuLP library [17].

### Practical applicability

Brazil’s flu sentinel network primarily requires sites in capital cities, with one site per 500,000 inhabitants, except in the South region, where one site is implemented for every 300,000 inhabitants, regardless of metropolitan status. Additionally, sample collection occurs in ICU services covering approximately 10% of the available ICU beds in each municipality, ensuring coverage across age groups [18]. The current network includes 310 sentinel sites in 199 municipalities, representing only 3.6% of the country’s cities.

To assess the practical applicability of our proposed sentinel network, we compared it with Brazil’s existing flu sentinel network to evaluate improvements in mobility and Indigenous region coverage. We also assessed whether the newly identified cities serve as key access points for healthcare services across different levels of complexity. This evaluation reinforces that the proposed sentinel locations align with urban centers that attract individuals seeking healthcare, making them strategic candidates for genomic sample collection.

## Results

To compose a more representative sentinel network for the country, we optimized the selection of N = 199 cities, the same number considered in Brazil’s flu sentinel network. In this network we achieved 100% DSEI coverage by selecting a total of 43.7% (87/199) cities that are fully inside of DSEI regions. In addition, 69.2% of the country’s mobility pattern is covered by these cities, that is 6,078,747 paths (of a total of 8,780,046 paths) originating in each city in the country.

In Brazil’s flu sentinel network, 53.8% (107/199) municipalities overlap 25 DSEIs (Figure 1 left panel). However, nine DSEIs lack any sentinel coverage: Altamira (Pará), Kayapó (Mato Grosso), Kayapó (Pará), Médio Rio Purus (Amazonas), Parque Indígena do Xingu (Mato Grosso), Rio Tapajós (Pará), Vale do Javari (Amazonas), Xavante (Mato Grosso), and Yanomami (Roraima). These regions span more than 750,515 km^2^ and are home to more than 1.11 million people. In addition, Brazil’s current flu sentinel network covers 52.4% (4,598,416 of 8,780,046 paths) of mobility patterns.

**Figure 1:**
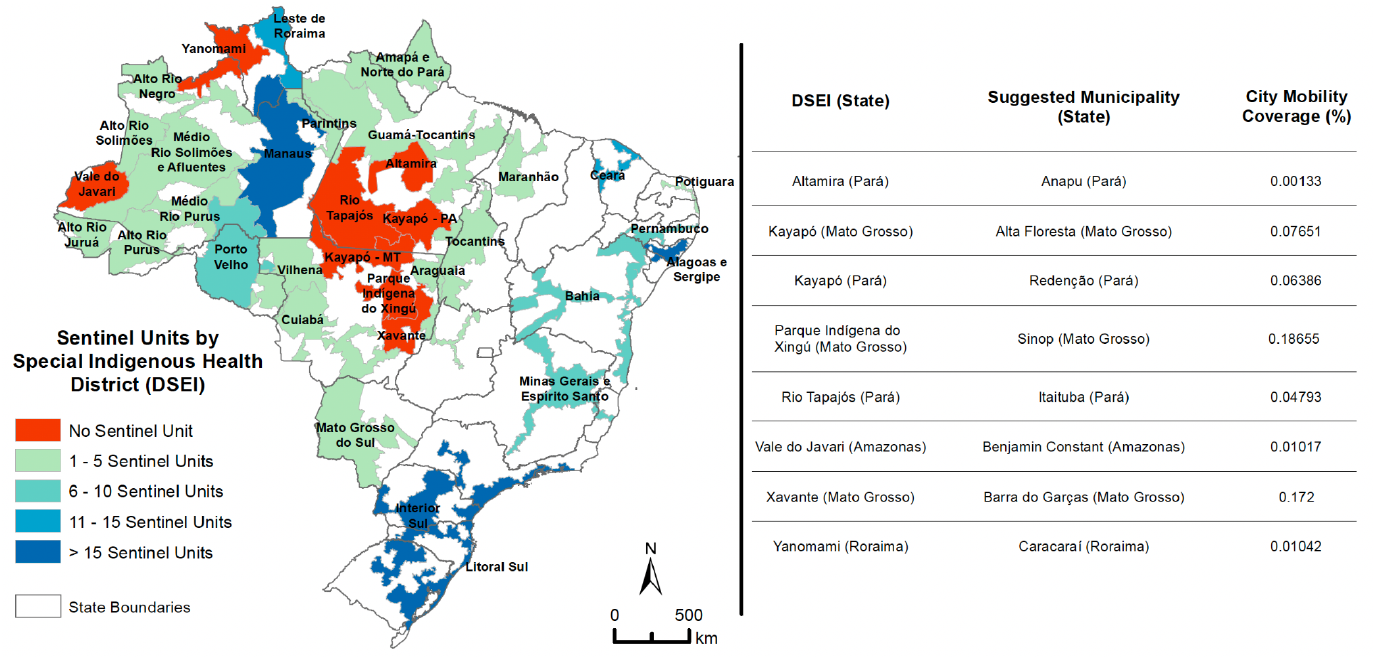
(Left) Current active sentinel distribution among Indigenous Special Health District. (Right) List of municipalities to guide the allocation of resources to improve early pathogen detection in under-served Indigenous regions, ensuring the enhancement of DSEI coverage while optimizing Brazilian mobility pattern coverage.

By rearranging 108 cities in Brazil’s flu sentinel network, keeping 91 cities already included in the current network, we achieved 100% DSEI coverage and increased national mobility coverage by 16.8 percentage points, from 52.4% to 69.2%. Figure 1 (right panel) shows the list of municipalities that now ensure coverage for DSEIs without sentinel sites in Brazil’s flu sentinel network.

Finally, we analyzed the movement of people seeking healthcare in the cities selected for the optimized sentinel network. None of the identified cities had a low healthcare attraction index. The majority (96%; 180 out of 199 cities) serve as hubs for high-complexity healthcare services, while 3% (7 out of 199) are classified as medium-complexity hubs. Among cities within DSEIs, 33.3% (29/87) have a high healthcare attraction index and are not currently covered by Brazil’s flu sentinel network. Additionally, 66.7% (58/87) have a medium attraction index, with four lacking sentinel coverage.

## Discussion

Our findings introduce a novel approach that integrates human mobility and Indigenous population coverage to optimally rearrange the current Brazil’s flu sentinel network locations for sample collection. By focusing on the historically marginalized Indigenous communities, the proposed approach aligns with expert recommendations to enhance surveillance in underserved areas [2-5,8], ultimately protecting vulnerable communities while improving Brazil’s ability to detect and respond to emerging health threats.

By ensuring that all DSEIs are represented by at least one sentinel site, we addressed surveillance gaps in nine previously uncovered regions, collectively home to over 1 million people. This outcome is particularly significant given the increased risks faced by Indigenous populations residing at the wildlife-urban interface. Our mobility-based optimization, combined with a restriction to 199 sentinel cities, reduced the number of cities within DSEIs compared to Brazil’s current flu sentinel network. This highlights two key considerations: (1) some DSEI regions covered in the existing network may be overserved, while (2) the total number of cities selected for sample collection may be insufficient to account for Brazil’s geographic and epidemiological diversity, potentially limiting early pathogen detection. Nevertheless, maintaining or expanding routine sentinel surveillance is paramount to establish a baseline of case incidence, as well as track circulating viruses [1], enabling the early detection of outbreaks, and the introduction of emergent pathogens. For instance, the absence of baseline testing likely contributed in delaying the recognition of the Zika virus outbreak in the Americas, which peaked in 2015-2016, but probably spread undetected since late 2013 [19, 20].

Furthermore, the optimized sentinel network achieved 69.2% mobility coverage, an improvement over Brazil’s current flu sentinel network. This shows that incorporating Indigenous population criteria resulted in only a slight decrease in mobility coverage compared to a network based solely on mobility patterns (70%, as shown in [8]). This underscores the need to examine what are the key epidemiological priorities, which may include socio-economic deprivation and regions with a high risk of climate-related disasters, to be taken into account when designing a representative sentinel network for early pathogen detection, as these factors may impact coverage metrics. Consequently, identifying an optimal balance between the minimum number of cities required for sentinel surveillance and the diversity of epidemiological priorities remains a critical area for future research.

Establishing a sentinel surveillance network, including a pipeline from sample collection to laboratory analysis, demands substantial resources, especially in regions with limited infrastructure. To address this challenge, we strategically centered our focus around DSEIs, an already in place health administrative structure in Brazil that provides primary care to Indigenous populations and articulates with other networks in the Unified Health System to guarantee access to medium- and high-complexity services. Additionally, by evaluating the health attraction index and taking into account the outdated requirement for sentinel network implementation established by the Brazilian Ministry of Health [18], we found that the newly selected cities already offer medium to high-complexity healthcare services, an indication that may support genomic sample collection. Leveraging this existing infrastructure not only facilitates routine surveillance but also strengthens overall healthcare capacity.

Our results contribute to increasing the likelihood of early pathogen detection without requiring an expansion in the number of sentinel sites, thereby addressing the critical challenge of limited funding—a well-documented barrier to improving global epidemiological surveillance, especially in low- and middle-income countries.

## Data Availability

All data produced are available online at GitHub directory Surveillance_Network_Indigenous_Mobility.

https://github.com/Julian-sun/Surveillance_Network_Indigenous_Mobility.git

## Author’s contributions

Conceptualization—JFO, MB-N, PIPR. Formal Analysis—ALA, AOV, JFO, MCLSC, PIPR. Writing (original draft, review and editing)—IM, JFO, MB-N, PIPR. Supervision—JFO, MB-N, PIPR. Project Administration—IM, MB-N, PIPR. Resources—MB-N.

## Acknowledgments

The findings reported are part of the Alert-Early System of Outbreaks with Pandemic Potential, an initiative under development by Brazil’s Fundação Oswaldo Cruz (Fiocruz) and the Federal University of Rio de Janeiro with financial support from the Rockefeller Foundation’s Health Initiative (grant 2023-PPI-007 awarded to M-BN). MB-N and PIPR are research fellows for Brazil’s National Council for Scientific and Technological Development (CNPq).

